# Toward Population-Level Brain AVM Screening: Construction and Evaluation of a Two-Step Genetic Risk Model Incorporating Rare Mendelian Variants and a Novel Polygenic Risk Score

**DOI:** 10.64898/2026.09.13.26362963

**Authors:** Pious D Patel, Sanjana Salwi, Penn Medicine Biobank, M. Reid Gooch, Adam Kundishora, Scott M. Damrauer, Stavropoula Tjoumakaris, Jan-Karl Burkhardt, Pascal Jabbour, Theodore G. Drivas

## Abstract

**Introduction:** Brain arteriovenous malformation (bAVM) is present in 0.01% of the population and has devastating neurologic consequences when ruptured. This study reports and evaluates a novel two-step genetic risk stratification model for bAVM, taking into account both rare Mendelian and common genetic variants, to identify patients with high genetic risk who can then receive screening MRI.

**Methods:** Participants in the Penn Medicine Biobank (PMBB) with both exome sequencing and imputed (TOPMED) genotyping array data were included in this study. Participants were stratified into a “Mendelian cohort” and a “non-Mendelian cohort.” Participants with a pathogenic or high probability loss-of-function variant in any known Mendelian AVM-related gene formed the Mendelian cohort, while the remainder formed the non-Mendelian cohort. To define bAVM cases, chart review was performed for all participants in the Mendelian group and for all other participants with at least one occurrence of the International Classification of Diseases (ICD)-10 code Q28.2 (“Arteriovenous malformation of cerebral vessels”) in their EHR. Within this reviewed group, those with MRI-, CTA-, or angiogram-confirmed brain AVMs were considered bAVM cases, those with negative imaging and all those participants lacking the ICD-10 Q28.2 code were considered controls. Those with ICD code Q28.2 but with inadequate imaging were excluded from analysis. A polygenic risk score (BAVM-PRS) was created using genetic data from a prior, external genome-wide association study of 1706 European-ancestry participants (515 bAVM cases, 1191 controls). Performance of the BAVM-PRS among non-Mendelian PMBB participants was assessed using logistic regression, density plots, receiver operating characteristics (ROC), and threshold-based analysis. Using this unbiased genotype-first approach to identify participants at high risk for bAVM, the prevalence of bAVM among both Mendelian risk and high-risk BAVM-PRS PMBB participants was calculated.

**Results:** A total of 56,332 participants from PMBB were included and stratified into a Mendelian cohort (n=86; median [interquartile range, IQR] age 55.6 [40.7, 64.5] years, 47.7% male, 67.4% European-ancestry, 30.2% African-ancestry) and a non-Mendelian cohort (n=56,246; median [IQR] age 56.4 [41.5, 66.8] years, 48.7% male, 73.9% European-ancestry, 21.4% African-ancestry). There were 4 (4.7%) confirmed bAVM cases in the Mendelian cohort and 55 (0.1%) in the non-Mendelian cohort. In the non-Mendelian cohort, BAVM-PRS Z-score was significantly higher among cases than controls (median [IQR] 0.303 [-0.218, 1.291] vs.-0.099 [-0.677, 0.566], p=0.002). Overall discrimination was moderate as assessed by ROC (Area-under-curve was 0.694, 95% CI 0.633-0.756). Density plots stratified by ancestry showed separation between cases and controls at high BAVM-PRS values for both European-and African-ancestry participants. Using a two-step risk stratification approach, flagging all Mendelian variant-carrying participants regardless of BAVM-PRS and all non-Mendelian participants with a BAVM-PRS Z-score >2.7 as high risk for bAVM, we identified a high risk population of 681 participants where prevalence of brain AVM was 1.0%. Within this high risk group, the number needed to screen (NNS) with brain MRI to diagnose one bAVM was 122, and our two-step risk stratification model demonstrated an overall capture rate of 12% with a sensitivity of 10% for bAVM in the PMBB population.

**Conclusion:** A two-step genetic risk model substantially improves bAVM screening efficiency, with a NNS of 122 to identify a single bAVM. Compared to a NNS of ∼1,250 when screening the general PMBB population, these sequential genetic filters represent a 10-fold increase in diagnostic efficiency. Similar approaches in other sequenced populations may allow for the efficient and early identification of bAVM in asymptomatic, high-risk individuals before they suffer the devastating sequelae of bAVM rupture.

## Introduction

Brain arteriovenous malformations (bAVM) are rare neurovascular lesions with severe neurologic consequences. There is an estimated prevalence of 10-18 cases per 100,000 persons and an annual intracranial rupture rate of 2-4%.^1–4^ Each intracranial rupture and subsequent hemorrhage has an approximately 20% rate of mortality and 45% rate of persistent functional impairment.^5,6^ Particularly devastating is that bAVM rupture is predominantly diagnosed in children and young adults, with 56% of diagnoses occurring before age 40.^6^

Despite these dire risks, there is no systematic screening framework for bAVM, largely owing to its rare disease status, multifactorial etiology, and lack of available biomarkers.^7–15^ While MRI brain has shown 80% sensitivity and 99% specificity for bAVM, global screening would require an estimated 12,500 MRI scans for one bAVM diagnosis.^16–18^

Several Mendelian genetic syndromes associated with high risk for bAVM have been identified. The hereditary hemorrhagic telangiectasia (HHT)-and capillary-malformation arteriovenous malformation syndrome (CM-AVM)-associated genes confer substantially increased risk for bAVM formation, with bAVM incidence in these patients reported to range from 2-13%.^19–21^ However, these bAVM prevalence estimates may be inaccurate; HHT and CM-AVM are both thought to be underdiagnosed,^22,23^ and most studies of HHT and CM-AVM have been completed using a phenotype-first approach, where patients with obvious features of the disease are ascertained for screening. A genotype-first approach, where all patients with pathogenic HHT and CM-AVM-associated genetic variants are screened for bAVM regardless of clinical features of disease, would provide a more accurate estimate of the true bAVM burden in these patient populations. However, no such study has been performed. Thus it is currently unknown how many individuals in the general population who carry these Mendelian variants might actually have a bAVM if they are otherwise clinically silent.

For non-Mendelian, sporadic, bAVMs, a previous genome-wide association study (GWAS) found an estimated heritability of 18% and identified several candidate single nucleotide polymorphisms (SNPs) significantly associated with sporadic bAVM diagnosis.^24^ Such foundational GWAS data can be used to calculate a polygenic risk scoring (PRS) to identify individuals at high polygenic genetic risk for bAVM. PRS is a method used to quantify genetic risk for a condition by summing the combinatorial effect of many low-effect-size SNPs across an individual. While PRS as a modality has shown variable performance on an individual patient level, it has demonstrated value on the population-scale to stratify groups of patients by risk for disease.^25^

The goal of this study was to construct and externally validate a novel, biologically informed polygenic risk score for brain arteriovenous malformation (BAVM-PRS), and to evaluate a two-step genetic risk model (Mendelian variant screening followed by polygenic risk scoring) to improve bAVM screening yield in a large, multi-ancestry biobank with rich genomic and phenotype data.

In the study, we first define the actual bAVM prevalence among Mendelian variant carriers using a genotype-first approach to address the diagnostic gap in clinically silent HHT/CM-AVM populations. Second, we construct and externally validate a novel, biologically informed polygenic risk score (BAVM-PRS) to quantify genetic risk and identify high-risk candidates for MRI screening within the non-Mendelian population. We then present the efficiency of a proposed two-step genetic risk model in guiding MRI-based screening for bAVM.

## Methods

### Population, Genotyping, and Sequencing

The Penn Medicine BioBank (PMBB) is a University of Pennsylvania academic biobank which recruits patient-participants from the University of Pennsylvania Health System around the greater Philadelphia area in the United States. Appropriate consent was obtained from each participant regarding storage of biological specimens, genetic sequencing, access to all available EHR data, and permission to recontact for future studies. The study was approved by the Institutional Review Board of the University of Pennsylvania and complied with the principles set out in the Declaration of Helsinki. This study included the subset of 56,332 individuals enrolled in PMBB who had previously undergone exome sequencing and genotyping array at the time of our study. Genotyping and exome sequencing had already been completed prior to our study.

Briefly, for each individual, DNA was extracted from stored buffy coats. Genotyping was performed by the Regeneron Genetics Center (Tarrytown, NY) as previously described.^26^ After performing sample-level quality control, genotype imputation was performed using Eagle v2.4.1 and Minimac4 version 1.0.0 software on the TOPMed Imputation Server. Imputation was performed for all autosomes, with TOPMed version R2 on a GRCh38 reference panel. Cosmopolitan post-imputation QC included imputation score filtering (R2 > 0.7), removal of palindromic variants, biallelic variant check, sex check, genotype call rate (>99%) and sample call rate (>99%) filtering, minor allele frequency filtering (MAF > 1%), and a Hardy–Weinberg equilibrium test (p-value > 1 × 10−8). From this data, PCAs were generated to allow for adjustment for population structure and to identify genetically informed ancestry (GIA) using EIGENSOFT version 7.2.0. Exome sequences were also generated by the Regeneron Genetics Center (Tarrytown, NY) as previously described and mapped to GRCh38.^26^ Sample-level filtering was as follows: individuals with low exome sequencing coverage (less than 75% of targeted bases achieving 20× coverage) or with high missingness (greater than 5% of targeted bases) were removed from analysis. Variant-level filtering was as follows: in each sample, all single nucleotide variants (SNVs) with a total read depth < 7 were changed to “no-call”, and similarly all insertion/deletion (INDEL) variants with a total read depth < 10 were changed to “no-call.”

### Stratification of Mendelian and Non-Mendelian Cohorts

To independently evaluate the effects of Mendelian risk and non-Mendelian, polygenic risk on bAVM prevalence, participants were stratified into a Mendelian cohort if they had a high-confidence predicted loss of function (pLoF) variant or known ClinVar-annotated pathogenic/likely pathogenic variant in any bAVM-associated gene (*ENG*, *ACVRL1*, *SMAD4*, *GDF2, RASA1,* and *EPHB4*). To define these variants, exome sequencing data were annotated using the Ensembl Variant Effect Predictor (VEP, version 102)^27^ with the plugin LOFTEE (version 0.3)^28^ to specifically annotate predicted loss of function (pLOF variants) and dbNSFP (version 4.2)^29^ to specifically annotate all single nucleotide variants. Only variants affecting NCBI RefSeq canonical transcripts were considered.^30^ Using this approach, 86 participants were identified as having a Mendelian bAVM risk and were analyzed as a separate cohort. The remaining participants formed the non-Mendelian cohort.

### Phenotyping

Patients with at least one occurrence of International Classification of Diseases (ICD) 10 code Q28.2 (“Arteriovenous malformation of cerebral vessels”) were identified as candidate cases of bAVM (n=176). All candidate cases, as well as all 86 patients in the Mendelian cohort, underwent manual chart and imaging review. True bAVM cases were defined by the presence of two strict criteria: 1) formal radiologic read of cerebral arteriovenous malformation, and 2) radiographic confirmation on MRI, CT angiography, or conventional angiography by reviewer or by attending neurosurgeon in a clinical note. Of the remaining ICD-10-code-positive candidates, those with negative imaging were considered controls (n=97), and those with no imaging to review were excluded from analysis (n=22). All remaining participants within the non-Mendelian cohort who did not have ICD code Q28.2 were classified as controls. There were a total of 57 validated ICD-10-identified bAVM cases, of which two were in the Mendelian cohort. After chart review of the Mendelian cohort, there were an additional n=2 bAVM cases identified which were not present in the ICD-10 candidate cases list. The final cohorts resulted in a total of 55 true bAVM cases and 56,191 bAVM controls in the non-Mendelian cohort, and four true bAVM cases and 82 bAVM controls in the Mendelian cohort.

### Polygenic Risk Score Construction

A polygenic risk score (BAVM-PRS) was constructed using summary statistics from a previously-performed GWAS of European-ancestry patients in the United States and Netherlands (515 cases, 1,191 controls).^24^ Recognizing that single-ancestry GWAS often fail to generalize in diverse cohorts, we developed a biologically-constrained PRS framework modeled after recent efforts to improve GWAS cross-population portability.^31,32^ Our framework mandated that every candidate SNP be both statistically significantly associated with bAVM in the GWAS and be assigned to a gene with a literature-backed role in one of eight bAVM-associated pathways (TGF-beta, Notch, VEGF, inflammatory, MAPK, vascular development, vascular endothelial growth, and Hedgehog) as annotated in the Weinsheimer *et al.* GWAS.^24^ This approach was designed to preserve the predictive value of the original GWAS summary statistics while filtering for a subset of SNPs with independently verified literature-backed evidence of gene-pathway involvement. By mandating this secondary functional requirement, we aimed to systematically remove the effect of ancestry-specific statistical anomalies – variants that may have reached significance due to population-specific linkage disequilibrium structures – in favor of markers representing causal biology which may be more conserved between populations. To empirically validate the utility of this biologically-constrained approach, we generated a traditional ‘all-SNP’ comparison PRS as well, allowing for a head-to-head assessment of our framework against standard, non-filtered genomic scoring.

For the primary BAVM-PRS, we extracted odds ratios and effect alleles for SNPs reaching a suggestive significance threshold (p<10^-4^) that were also annotated to genes within one of the eight aforementioned bAVM-related signaling pathways in the original GWAS.^24^ These SNPs were matched to the format of the PMBB imputed genotype dataset after deriving chromosome, position, and alleles from the GRCh38.p14 Primary Assembly. The BAVM-PRS was then generated using direct scoring from the converted beta value effect sizes for a total of 27 SNPs. The score was applied for each participant using PLINK2 (v2.0)^33^ to compute the sum of the number of effect alleles at each locus weighted by the published beta value across all selected SNPs (**Table S1**). The resulting raw PRS values were subsequently standardized within the entire population using Z-score transformation (mean=0, standard deviation =1). Reweighting or PRS-fitting was specifically not performed in order to keep the external validation completely independent.

An additional comparison PRS, BAVM-PRS-All, was created using direct scoring from the reported effect sizes of all SNPs reported in the GWAS performed by Weinsheimer et al., regardless of gene annotation. All 57 SNPs identified in the original discovery GWAS as significantly associated with bAVM were mapped to the GRCh38.p14 Primary Assembly.^24^ The PRS was then constructed identically to BAVM-PRS except that it included this full list of SNPs instead of only the 27 biologically-plausible ones. The full components of this score are in **Table S1**.

### Analysis

For the non-Mendelian cohort, Wilcoxon rank sum test and univariate logistic regression were performed with BAVM-PRS modeled as a continuous variable to measure the association between BAVM-PRS and bAVM diagnosis. Receiver operating characteristics (ROC) and area-under-curve (AUC) were calculated using BAVM-PRS score alone as well as with covariate adjustment (age at time of PMBB enrollment, sex, and principal components (PCs) 1-6 of genomic data). Density plots were generated to illustrate the distribution of BAVM-PRS values between bAVM cases and controls within this non-Mendelian population. All analyses performed for the primary BAVM-PRS were replicated and compared for the all-inclusive, BAVM-PRS-All.

Threshold values were determined through a combination of the Youden index^34^ and optimization for prevalence. To standardize our threshold-based risk stratification and ensure reproducibility, we mapped our findings onto the risk-modeling evaluation framework proposed by Pfeiffer and Gail (2011).^35^ Specifically, we converted our clinical score cutoffs into two standardized epidemiological metrics: the Proportion of Cases Followed (PCF) and the Proportion Needed to Follow (PNF). The PCF(q) represents the proportion of total cases captured when screening the top q fraction of the population at highest genetic risk. PNF(p) indicates what proportion of the highest-risk population must be followed to capture a predefined proportion p of total cases. Capture rate (which we defined as equivalent to PCF) was defined as the number of cases identified in the high-risk subgroup divided by the number of cases in the whole population. Prevalence was reported as the number of cases in the high-risk subgroup divided by the total number of participants in the high-risk subgroup. NNS was calculated as the inverse of prevalence multiplied by sensitivity of imaging, assuming that screening brain MRI has 80% sensitivity for bAVM diagnosis.^16^ Specifically, two thresholds were selected from the non-Mendelian cohort: one representing a balance of capture rate and prevalence and a more stringent threshold optimized for maximal prevalence.

For the Mendelian cohort, summary statistics and bAVM prevalence were reported for the whole subpopulation and stratified by Mendelian disease gene. NNS was identically calculated for this cohort based on the baseline bAVM prevalence. In addition, density plots and median PRS scores were generated to visualize the performance of BAVM-PRS in the Mendelian population to evaluate if polygenic risk provided further case stratification.

### Ancestry-stratified Analysis

The BAVM-PRS we created was derived from a European-ancestry-only GWAS cohort. To test the performance of the BAVM-PRS across patients of different genetically informed ancestry (GIA) groups, an ancestry-stratified analysis was performed. One subset population was created for participants with European GIA and one for those with African GIA, the two largest GIA groups in the PMBB cohort. The analysis was repeated within the non-Mendelian subsets using cohort-specific Z-scores, which were recalculated from the raw BAVM-PRS to ensure accurate normalization within each ancestry group. To evaluate the portability of the scoring system, the thresholds derived from the non-Mendelian cohort were applied to each subpopulation; additional ancestry-specific thresholds were then identified to optimize screening performance based on each cohort’s unique score distribution.

### Construction of the Two-step Genetic Screening Model

We constructed a two-step genetic risk stratification model to maximize capture rate alongside screening efficiency. The first step screens the population for rare, Mendelian predicted pathogenic variants in *ENG*, *ACVRL1*, *SMAD4*, *GDF2, RASA1,* or *EPHB4*. Any individual carrying a high-confidence pLoF or an annotated pathogenic/likely pathogenic in any one of these genes is directed toward brain MRI screening. Individuals who do not carry a Mendelian risk variant proceed to the second tier of the model. For this second tier, a standardized BAVM-PRS polygenic risk score is calculated and converted to a population-wide Z score. Individuals exceeding the stringent threshold BAVM-PRS score (described in *Analysis* section) are directed toward brain MRI. Individuals who do not carry a Mendelian risk variant and who do not exceed the stringent BAVM-PRS threshold are not screened.

## Results

### Population Characteristics and bAVM Prevalence

A total of 56,332 participants from the PMBB were included and stratified into a Mendelian cohort (n=86; median [IQR] age 55.6 [40.7, 64.5] years, 47.7% male, 67.4% European-ancestry, 30.2% African-ancestry) and a non-Mendelian cohort (n=56,246; median [IQR] age 56.4 [41.5, 66.8] years, 48.7% male, 73.9% European-ancestry, 21.4% African-ancestry) (**Table 1**).

**Table 1:** Baseline Characteristics of the Study Population.

|  | Non-Mendelian Cohort | Mendelian Cohort |
| --- | --- | --- |
| Cohort Size | 56,246 | 86 |
| Confirmed Brain Arteriovenous Malformation Cases, n (%) | 55 (0.10%) | 4 (4.7%) |
| Male Sex, n (%) | 27,375 (48.7%) | 41 (47.7%) |
| Age in years, median [IQR] | 56.4 [41.5, 66.8] | 55.6 [40.7, 64.5] |
| European Ancestry, n (%) | 41,575 (73.9%) | 58 (67.4%) |
| African Ancestry, n (%) | 12,009 (21.4%) | 26 (30.2%) |
| Other Ancestry, n (%) | 2,662 (4.7%) | 2 (2.3%) |
| BAVM-PRS, median [IQR] |  |  |
| – Entire Cohort | -0.098 [-0.677, 0.567] | -0.095 [-0.629, 0.950] |
| – Cases | 0.303 [-0.218, 1.291] | -0.159 [-0.667, 0.369] |
| – Controls | -0.099 [-0.677, 0.566] | -0.095 [-0.629, 1.042] |
| p-value | 0.002 | 0.697 |

Summary of demographic and genetic characteristics for the Mendelian (high confidence pLOF and ClinVar pathogenic/likely pathogenic variants in *ENG*, *ACVRL1*, *SMAD4*, *GDF2*, *RASA1*, or *EPHB4*) and non-Mendelian (all other individuals) cohorts. AVM-PRS values are presented as Z-scores standardized across the whole population. P-values represent the results of Wilcoxon rank sum test comparing PRS distributions between bAVM cases and controls within each respective cohort. *IQR*, *Interquartile Range*; *bAVM*: *Brain Arteriovenous Malformation*; *PRS*: *Polygenic Risk Score*.

In the non-Mendelian cohort, a total of 55 true bAVM cases were identified, corresponding to a PMBB population prevalence of 0.1% (55/56,246). A total of 56,191 participants were classified as controls. In the Mendelian cohort, 50 participants had sufficient imaging to rule in or out the presence of a bAVM. A total of 4 out of 50 participants had confirmed bAVM, corresponding to a prevalence of 8% among Mendelian cohort participants with brain imaging (3/10 *ENG*, 1/18 *ACVRL1*, 0/3 *SMAD4*, 0/5 *GDF2*, 0/2 *RASA1*, 0/12 *EPHB4*). Considering all participants in the Mendelian cohort, including those without brain imaging, a total of 4 out of 86 participants had confirmed bAVM, corresponding to a prevalence of 4.7% (3/16 *ENG*, 1/32 *ACVRL1*, 0/4 *SMAD4*, 0/9 *GDF2*, 0/5 *RASA1*, 0/20 *EPHB4*). Based on this 4.7% prevalence and assuming an 80% sensitivity for brain MRI, the estimated number needed to screen (NNS) for this Mendelian cohort is no more than 27 with a capture rate of at least 7% (4/59) of total bAVM cases in the overall PMBB population (**Table 2**).

**Table 2:** Screening Performance of the Two-Step Genetic Risk Model Compared to Established Clinical Screening Benchmarks.

| Condition | Screening Modality | Risk Enrichment Approach | Capture Rate | Sensitivity | Prevalence | Number needed to screen per diagnosis |
| --- | --- | --- | --- | --- | --- | --- |
| Brain arteriovenous malformation | Brain MRI | **Two-step Genetic Risk Model** | **12% (7/59)** | **10%** | **1.0% (7/681)** | **122** |
|  |  | Mendelian Risk Variant alone | 7% (4/59) | 6% | 4.7% (4/86) | 27 |
|  |  | BAVM-PRS > 2.7 alone | 5% (3/59) | 4% | 0.50% (3/595) | 248 |
| Breast cancer<sup>36</sup> | Mammography | Women of age > 40 years | — | 87% | 0.51% (8,529/1,682,504) | 197 |
| Lung cancer<sup>37,38</sup> | Chest CT | Adults of age 50-80 years with ≥20 pack-years | — | 97%<sup>38</sup> | 0.90% (203/22,600)<sup>37</sup> | 115 |
| Developmental dysplasia of the hip<sup>39,40</sup> | Physical exam ± ultrasound | Infants | — | 57%<sup>40</sup> | 1.40%*<sup>39</sup> | 125 |

A comparison of the number needed to screen, capture rate, sensitivity, and prevalence for different conditions and their recommended screening modalities. For the bAVM screening paradigm described in this manuscript, values for a rare Mendelian variant alone, BAVM-PRS alone, and a "two-step model" are shown. The two-step model combines identification of all high risk Mendelian variant carriers with a high-risk non-Mendelian cohort defined by BAVM-PRS threshold of >2.7. For bAVM screening: capture rate was defined as the proportion of total bAVM cases identified by the specified risk enrichment approach; sensitivity was calculated as the product of the capture rate and the established 80% sensitivity of brain MRI for bAVM detection; prevalence was defined as the disease frequency specifically within the subset of patients meeting the listed risk enrichment criteria (e.g., the number of patients with bAVM among the 681 individuals identified as high-risk by the Two-Step Model); number needed to screen was calculated as the inverse of (Prevalence × Imaging Sensitivity). Benchmarks for breast cancer, lung cancer, and developmental dysplasia of the hip are derived from published screening outcomes (\*prevalence for DDH derived from pooled meta-analysis study).<sup>36-40</sup>

### Diagnostic Association and Screening Performance of BAVM-PRS in the Non-Mendelian Cohort

We constructed a BAVM-PRS as described in the Methods section to attempt to stratify the overall, non-Mendelian PMBB population by bAVM polygenic risk. In the non-Mendelian cohort, there was a significant difference in BAVM-PRS values (defined as Z-score within the entire cohort) between AVM cases and controls (median [IQR] 0.303 [-0.218, 1.291] vs.-0.099 [-0.677, 0.566], p=0.002) (**Figure 1**). Unadjusted logistic regression showed that BAVM-PRS alone was significantly associated with bAVM diagnosis (OR=1.50, 95% CI 1.18-1.90, p<0.001).

**Figure 1:**
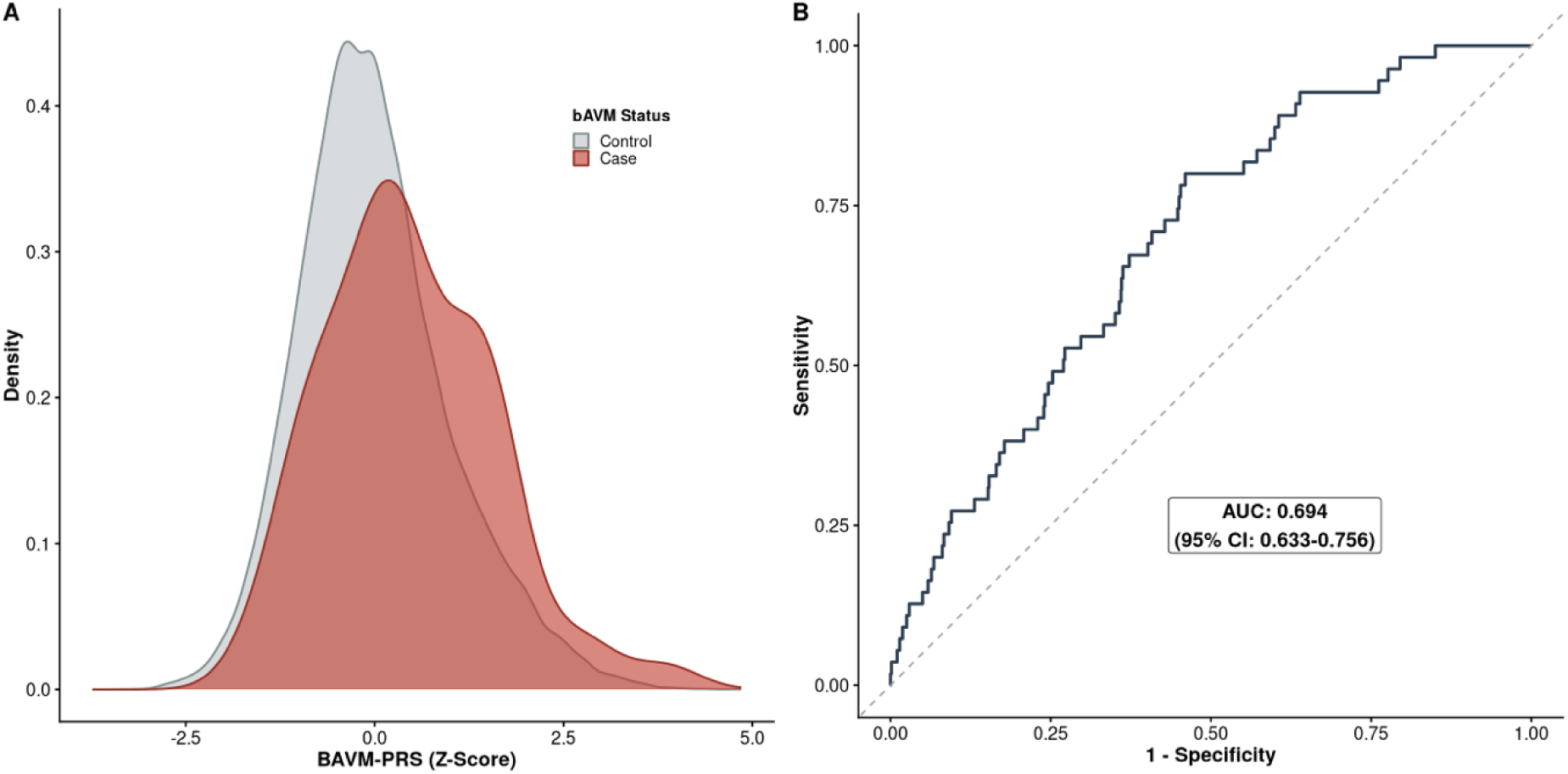
Distribution and Diagnostic Performance of the BAVM-PRS in the Non-Mendelian Cohort. (A) Density Distribution of BAVM-PRS scores (represented as Z-score) is shown for confirmed bAVM cases (red, n = 55) and controls (grey, n = 56,191) within the non-Mendelian cohort. There is a significant shift in the median PRS for cases compared to controls (median [IQR] 0.303 [-0.218, 1.291] vs.-0.099 [-0.677, 0.566], p = 0.002). (B) Receiver operating characteristic (ROC) curve illustrating the discriminatory performance of BAVM-PRS for identifying brain arteriovenous malformations in the non-Mendelian PMBB population. The BAVM-PRS achieved an area under the curve (AUC) of 0.694 (95% CI: 0.633–0.756), representing modest but statistically significant discrimination. Logistic regression confirmed that BAVM-PRS as a univariate continuous variable is significantly associated with bAVM diagnosis (OR = 1.50 per SD increase, 95% CI: 1.18–1.90, *p* < 0.001). Abbreviations: BAVM, brain arteriovenous malformation; CI, confidence interval; PMBB, Penn Medicine BioBank; PRS, polygenic risk score; IQR, Interquartile Range.

Density plot revealed notable separation of BAVM-PRS scores between bAVM cases and controls, and ROC analysis of BAVM-PRS, adjusted for age, sex, and PCs 1-6, showed modest discrimination (AUC 0.694, 95% CI 0.633-0.756) (**Figure 1**). Threshold-based analysis revealed increasing prevalence of bAVM with increasing BAVM-PRS thresholds. At a threshold of BAVM-PRS > 1.1, representing the top 13.7% of the population at highest genetic risk (q=0.137), the capture rate, equivalent to Proportion of Cases Followed (PCF), was 29% (17/59 of total cases). This corresponds to a Proportion Needed to Follow (PNF) of 13.7% to achieve this level of case stratification.^35^ At this threshold of BAVM-PRS > 1.1, prevalence was 0.22% (17/7,720), providing an estimated number needed to screen of 568 brain MRIs for one bAVM diagnosis (with assumption of 80% sensitivity from MRI).^16^ At a more stringent threshold of BAVM-PRS > 2.7, representing the top 1.06% of the population at highest risk (q=0.0106), the model achieved a capture rate of 5% (3/59 of total cases), meaning a PNF of only 1.06% is required to capture these ultra-high-risk individuals. At this cutoff, prevalence was 0.50% (3/595), providing NNS of 248. Compared to an estimated population-level NNS of approximately 1,250, in PMBB this represents a 5-fold improvement in screening efficiency (**Table 2**).

### Ancestry-stratified Diagnostic Association and Screening Performance of BAVM-PRS

A total of 41,575 non-Mendelian participants were of European GIA (n=29 with bAVM diagnosis, median [IQR] age 58.0 [43.9, 67.9] years, 52.3% male) and 12,009 were of African GIA (n=21 with bAVM diagnosis, median [IQR] age 53.2 [38.6, 63.4] years, 36.9% male). BAVM-PRS distribution stratified by ancestry is shown in **Figure 2** and statistical association shown in **Table 3**.

**Figure 2:**
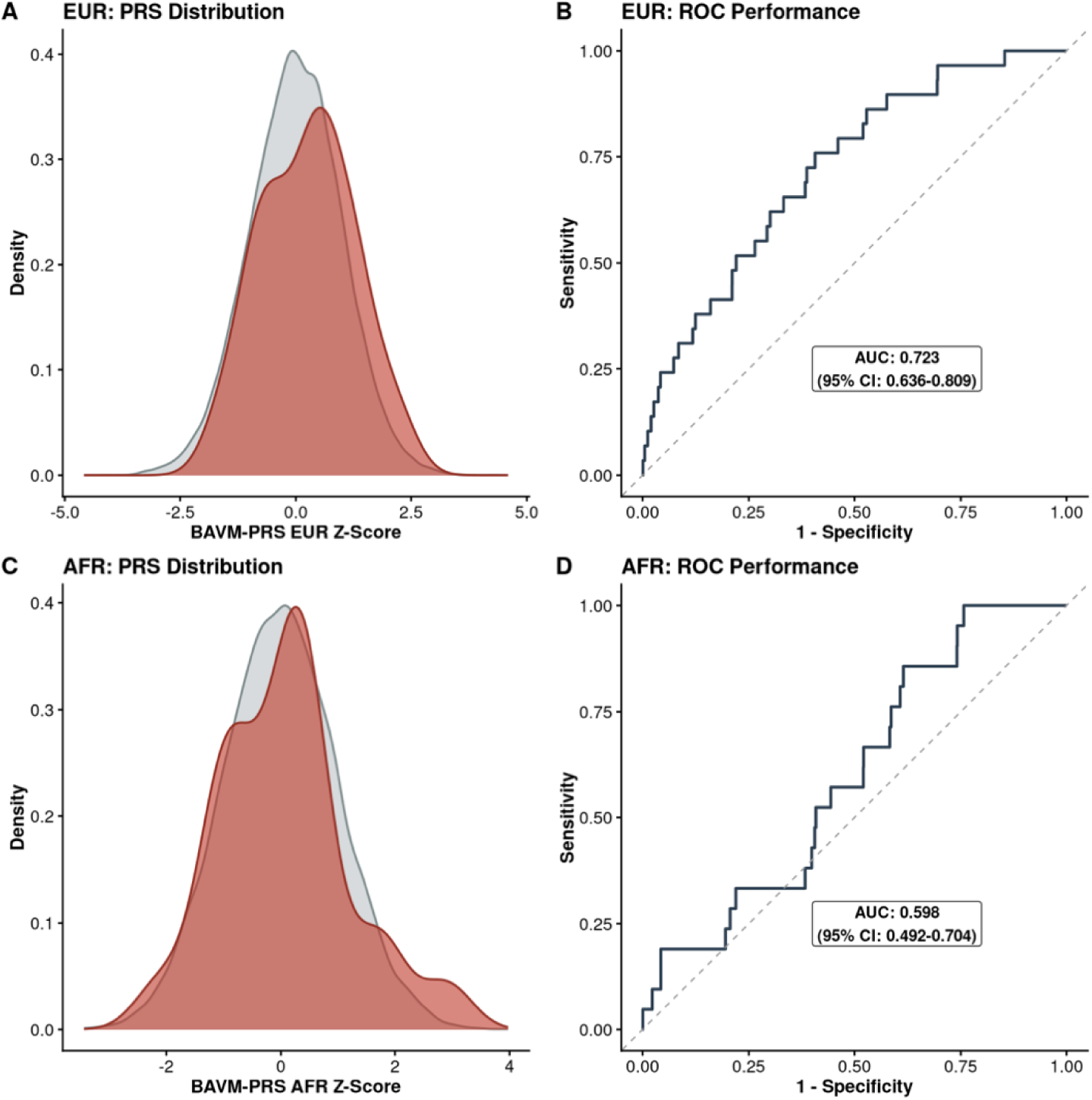
Ancestry-Stratified Performance of the BAVM-PRS in the Non-Mendelian Cohort. (A) Density Distribution of BAVM-PRS scores (represented as Z-score) is shown for confirmed bAVM cases (red, n = 29) and controls (grey, n = 41,546) within the non-Mendelian cohort specifically for individuals of European GIA. There is a nonsignificant trend towards the median PRS being higher for cases compared to controls (median [IQR] 0.361 [-0.635, 0.897] vs. 0.010 [-0.662, 0.671], p = 0.186). (B) Receiver operating characteristic (ROC) curve illustrating the discriminatory performance of BAVM-PRS for identifying brain arteriovenous malformations in the non-Mendelian PMBB population for individuals of European GIA. The BAVM-PRS achieved an area under the curve (AUC) of 0.723 (95% CI: 0.636–0.809), representing modest but statistically significant discriminating ability. (C) Density Distribution of BAVM-PRS scores (represented as Z-score) is shown for confirmed bAVM cases (red, n = 21) and controls (grey, n = 11,988) within the non-Mendelian cohort specifically for individuals of African GIA. There is a nonsignificant trend towards the median PRS being higher for cases compared to controls (median [IQR] 0.189 [-0.761, 0.452] vs.-0.002 [-0.675, 0.672], p = 0.962). (D) Receiver operating characteristic (ROC) curve illustrating the discriminatory performance of BAVM-PRS for identifying brain arteriovenous malformations in the non-Mendelian PMBB population for individuals of African GIA. The BAVM-PRS achieved an AUC of 0.598 (95% CI: 0.492–0.704). Abbreviations: AFR, African GIA; AUC, area under the curve; BAVM, brain arteriovenous malformation; CI, confidence interval; EUR, European GIA; PRS, polygenic risk score; IQR, Interquartile Range.

**Table 3:** Comparative Performance of the Biologically-Constrained BAVM-PRS versus the All-SNP BAVM-PRS-All Across Ancestry Groups.

| PRS Z Score | Ancestry Cohort | Number bAVM Cases | AUC | Median [IQR] PRS Z-score for bAVM Cases | Median [IQR] PRS Z-score for Controls | P-value |
| --- | --- | --- | --- | --- | --- | --- |
| **BAVM-PRS (biologically-relevant SNPs only)** | All participants (*n*=56,246) | 55 | 0.694 | 0.303 [-0.218, 1.291] | -0.099 [-0.677, 0.566] | 0.002 |
|  | European ancestry (*n*=41,575) | 29 | 0.723 | 0.361 [-0.635, 0.897] | 0.010 [-0.662, 0.671] | 0.186 |
|  | African ancestry (*n*=12,009) | 21 | 0.598 | 0.189 [-0.761, 0.452] | -0.002 [-0.675, 0.672] | 0.962 |
| **BAVM-PRS-All (all SNPs from Weinsheimer *et al.*)** | All participants (*n*=56,246) | 55 | 0.682 | 0.072 [-0.643, 0.905] | 0.107 [-0.619, 0.681] | 0.613 |
|  | European ancestry (*n*=41,575) | 29 | 0.700 | -0.145 [-0.613, 0.627] | 0.146 [-0.608, 0.711] | 0.382 |
|  | African ancestry (*n*=12,009) | 21 | 0.592 | 0.314 [-0.790, 0.724] | 0.042 [-0.680, 0.692] | 0.724 |

Comparison of area under the receiver operating characteristic curve (AUC) and median [IQR] standardized score between the BAVM-PRS and BAVM-PRS-All PRS constructions in the non-Mendelian cohort. Results are shown for the entire population, and stratified by genetically informed ancestry (GIA) for the European ancestry and African ancestry cohorts. PRS values represent the Z-score recalculated within that specific ancestry cohort. P-values are the result of a Wilcoxon rank sum test between PRS scores for bAVM cases and controls.
*SNP: Single Nucleotide Polymorphism; AUC: Area Under the Curve; IQR, Interquartile Range.*

Among European-ancestry participants, the BAVM-PRS demonstrated an AUC of 0.723 (95% CI 0.636-0.809). Utilizing entire-population Z-scores at the previously defined thresholds, a score >1.1 yielded a capture rate of 24% (7/29) and a prevalence of 0.12% (7/5,453), while at >2.7, capture rate and prevalence both fell to 0% (0/29 and 0/152). Optimization of screening thresholds for the European GIA subpopulation revealed that a BAVM-PRS threshold of >2.0 identified a high-risk group with a capture rate of 7% (2/29) and a prevalence of 0.22% (2/895), representing a 3.1-fold increase over the European GIA cohort’s overall bAVM prevalence (0.070%).

In the African-ancestry cohort, the BAVM-PRS achieved an AUC of 0.598 (95% CI 0.492-0.704). Utilizing entire-population Z-scores, a threshold of >1.1 demonstrated a capture rate of 14% (3/21) with a prevalence of 0.19% (3/1,589), and a threshold of >2.7 yielded a capture rate of 5% (1/21) and a prevalence of 2.5% (1/40). Optimization of screening thresholds for the African GIA subpopulation showed that a BAVM-PRS threshold of >2.8 identified a high-risk cohort with a capture rate of 5% (1/21) and a prevalence of 3.03% (1/33), representing a 17.8-fold improvement in screening efficiency compared to the African GIA cohort’s overall bAVM prevalence (0.17%).

### Comparison between the Biologically-Restricted BAVM-PRS and BAVM-PRS-All

BAVM-PRS-All, the PRS constructed from all significant SNPs reported in Weinsheimer *et al.,* regardless of gene annotation, showed modest discriminatory ability (**Figure S1**, AUC 0.682, 95% CI 0.616-0.747, adjusted for age, sex, and PCs 1-6), comparable to that of the biologically informed BAVM-PRS (**Figure 1**), but lacked a significant or consistent directional association with bAVM status. **Table 3** reports the performance of the biologically informed BAVM-PRS compared to BAVM-PRS-All. Specifically, for BAVM-PRS-All, the standardized score for bAVM cases (median [IQR] 0.072 [-0.643, 0.905]) was not significantly elevated compared to controls (median [IQR] 0.107 [-0.619, 0.681], p=0.613). There was, additionally, a bimodal distribution of BAVM-PRS-All scores among bAVM cases (**Figure S1**) which was notable in both European GIA and African GIA subpopulations (**Figure S2**).

### Performance of BAVM-PRS Within the Mendelian Cohort

While BAVM-PRS showed significant separation between cases and controls in the non-Mendelian cohort, it showed no separation between cases and controls in the Mendelian cohort (**Figure 3**). Median [IQR] BAVM-PRS scores were-0.159 [-0.667, 0.369] for Mendelian cases (n=4) and-0.095 [-0.629, 1.042] for Mendelian controls (n=82) with no significant statistical difference between the two groups (p=0.697). However, these comparative analyses are notably underpowered due to the limited number of bAVM cases in this group.

**Figure 3:**
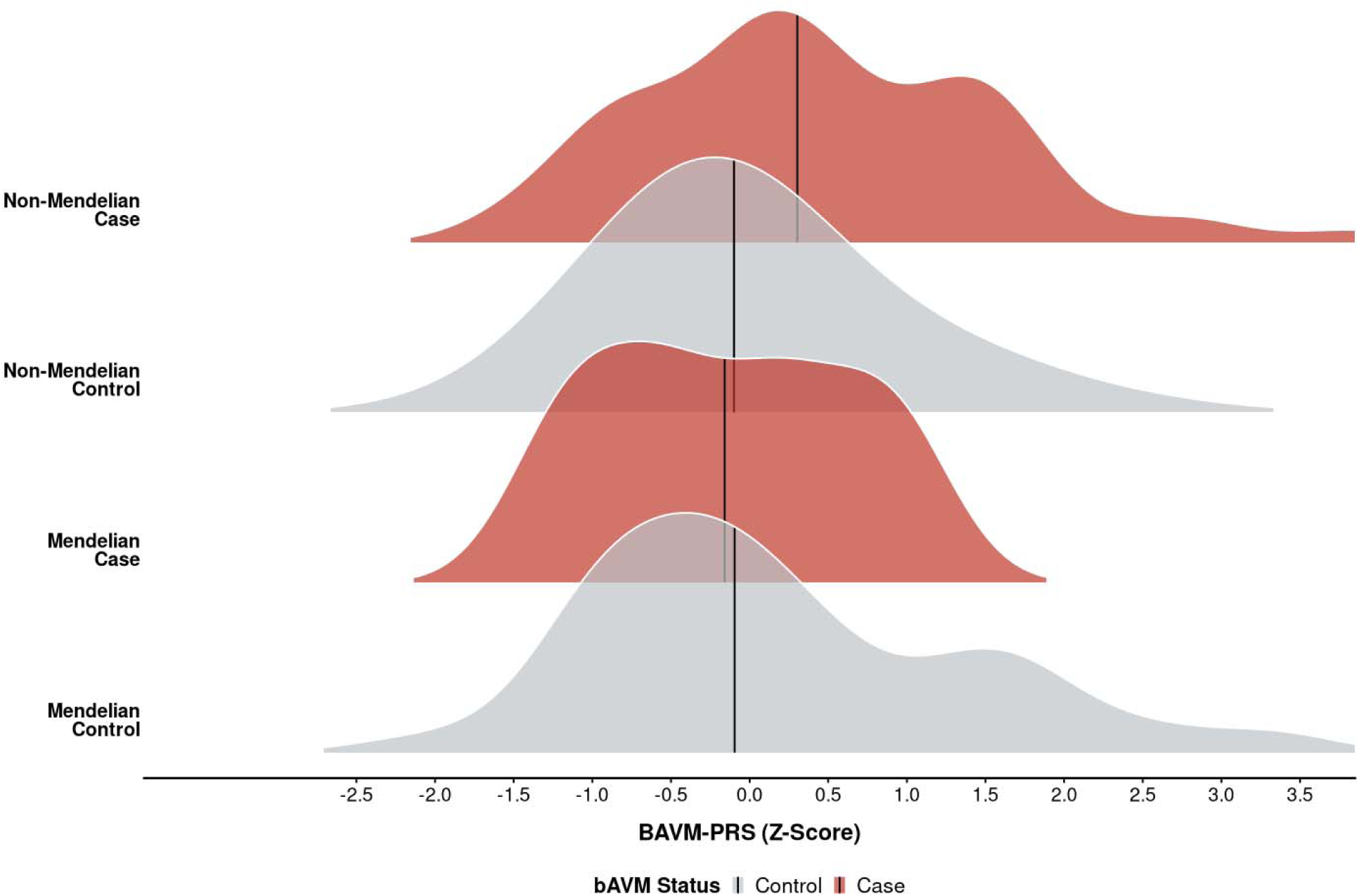
Comparative Performance of the BAVM-PRS in Mendelian versus Non-Mendelian Cohorts. Ridge plot illustrates the distribution of the BAVM-PRS (represented as a Z-score) across cases and controls between the non-Mendelian and Mendelian groups. Vertical black lines indicate the median PRS value for each group. The Mendelian group (4 bAVM cases, 82 controls) represents the cohort of individuals with high-confidence predicted loss-of-function (pLoF) or known pathogenic/likely pathogenic variants in established bAVM-associated genes (*ENG, ACVRL1, SMAD4, GDF2, RASA1,* and *EPHB4*). The non-Mendelian group (55 bAVM cases, 56,191 controls) represents the study cohort, excluding individuals with these high-confidence rare variants. While the BAVM-PRS in non-Mendelian cases is shifted higher compared to non-Mendelian controls, BAVM-PRS demonstrates no discriminatory power in the Mendelian group. The lack of separation in the Mendelian group suggests that in the presence of highly penetrant rare variants, the cumulative effect of common polygenic variants – as captured by the BAVM-PRS – may not significantly affect overall bAVM risk. Abbreviations: bAVM, brain arteriovenous malformation; BAVM-PRS, brain arteriovenous malformation polygenic risk score; IQR, interquartile range; pLoF, predicted loss-of-function; PRS, polygenic risk score.

### Performance of Two-step Genetic Screening Model

The performance of the two-step genetic risk framework (first identifying Mendelian variant carriers for bAVM screening, with subsequent screening of non-Mendelian variant carriers with a BAVM-PRS score above a stringent cutoff of >2.7) was assessed using capture rate and prevalence. This framework identified a combined cohort of 681 bAVM-high-risk-individuals for brain MRI screening (86 with Mendelian risk, 595 with BAVM-PRS >2.7). Within this cohort, bAVM prevalence is 1.0% (7 bAVM cases total; 4 in the Mendelian cohort, 3 in the high PRS cohort). This corresponds to a capture rate of 12% (7 bAVM cases identified from 59 total cases in the overall PMBB population). On a population scale, this two-step model delivers a number needed to screen of 122, representing a 10-fold improvement compared to the unstratified PMBB baseline of NNS of 1,250 (overall PMBB bAVM prevalence of 0.1%), assuming 80% sensitivity of screening brain MRI.

## Discussion

The consequences of untreated bAVM are devastating, with a ∼80% lifetime risk of rupture when diagnosed by early adulthood.^3,4^ However, the low overall prevalence precludes standard MRI screening for the general population,^1^ which would require an estimated number needed to screen of 1,250 patients for one bAVM diagnosis (using the overall prevalence of 0.1% in the PMBB population studied here).^16^ Here we have developed and evaluated a two-step genetic model for identifying individuals at high risk for bAVM using a combination of rare Mendelian pathogenic variant identification and a novel polygenic risk score for bAVM in the non-Mendelian population. Each step of our model identifies high risk bAVM individuals using different modalities. Our analyses showed that, while rare (present in only 0.15% of the examined population), the presence of a pathogenic Mendelian bAVM variant was associated with at least a 4.7% prevalence of bAVM. At the same time, in individuals without rare bAVM-associated Mendelian variants, the prevalence of bAVM was only 0.1%. However, in this population, higher BAVM-PRS score was significantly associated with a higher likelihood of bAVM diagnosis. Combining both approaches, we show that it is possible to identify a set of individuals within a population at high risk for bAVM, where the number needed to screen with brain MRI is only 122 to identify a single bAVM case, with 12% of all bAVM cases ascertained. Importantly, our analyses were performed in an ancestry-diverse biobank population and show promise for being generalizable across ancestry groups.

This number needed to screen (NNS) of 122 to identify a single bAVM for our two-step model is comparable to the NNS per diagnosis for other accepted population screening models: mammography for women > age 40 has a NNS of 197 per breast cancer diagnosis;^36^ chest computed tomography scans in patients 50-80 years old with a >=20 pack year smoking history has a NNS of 115 per lung cancer diagnosis;^37,38^ physical exams and selective ultrasonography has a NNS of 125 to identify a single infant with developmental dysplasia of the hip (DDH).^39,40^ However, while the NNS for our proposed two-step genetic risk model is similar to those of these accepted screening guidelines, the sensitivity is notably lower – 10% for our two-step bAVM screening paradigm, compared to 57-97% for breast cancer, lung cancer, and DDH (**Table 2**). This is a significant limitation of our approach, using solely germline genetic risk. This limitation is mirrored in the modest AUC (0.694) we find for BAVM-PRS within the non-Mendelian population. However, in the context of a rare disease where initial rupture can be fatal or permanently disabling, we argue that even 10% sensitivity provides substantial clinical value by identifying individuals who would otherwise never receive screening. Through this lens, a 10% sensitivity represents an opportunity to prevent 10% of the catastrophic neurologic events associated with bAVM, while limiting the number of individuals screened to only those of high genetic risk.

At the same time, the results of our study also highlight how germline genetic risk alone is likely not sufficient to predict bAVM in all patients. The majority of patients with either high BAVM-PRS or with a Mendelian bAVM risk variant were found to be unaffected by bAVM; simultaneously, the majority of bAVM cases occurred in individuals outside of the high risk group that we are able to define using germline rare and common genetic variation. This speaks to the putative model for bAVM formation which involves both germline genetic predisposition and environmental “second hits.”^7^ Brain AVMs have been strongly associated with low allele frequencies somatic activating mutations in *KRAS* leading to dysregulation in the MAPK pathway.^8^ These RAS pathway alterations, in addition to the previously described roles of increased VEGF and Notch activity, appear to play an endothelium-specific role in the early development of these malformations. ^9–13^ Additionally, other non-genetic instigators of bAVM formation, including disordered inflammatory and immune response, breakdown of the blood-brain barrier, endothelial migration, and recruitment of smooth muscle cells, have been reported.^14,15^ Thus, strategies to improve the sensitivity of the germline genetic screening paradigm we propose here would likely require combining this study’s Mendelian and BAVM-PRS genetic risk model with additional clinical risk factors (e.g. cell-free DNA sequencing, serum/CSF markers, vessel wall imaging) that capture the somatic and environmental triggers of bAVM formation, but which are also significantly more difficult to obtain at a population scale.

Exactly how and when to implement a bAVM screening program like the one we propose here is a matter of discussion. As the world of newborn screening moves from metabolic testing to population-level whole genome sequencing in infancy,^41^ one could envision implementing such a bAVM genetic risk model into newborn screening approaches. Newborn screening panels are designed to identify rare, treatable disorders which would have dire consequences from missed diagnosis (e.g. cystic fibrosis, phenylketonuria, spinal muscular atrophy). Mendelian and polygenic risk-driven bAVM screening would parallel this goal, given the disease’s 2-4% annual risk of rupture and successful treatment rates of up to 95% for small lesions.^4,42^ Alternative approaches could utilize large-scale, “opt-in” sequenced biobanks similar to PMBB. This approach would benefit from the integration of real-world clinical data into risk assessments while preserving patient autonomy regarding genetic data use. However, this approach is limited by ascertainment bias, allowing inclusion of only those individuals who actively engage with the health system. Moreover, applying a bAVM screening program in adulthood misses the critical childhood and early-adulthood window during which a substantial proportion of bAVM ruptures occur.^6^

In any context, the clinical implementation of a bAVM screening framework carries significant ethical and practical considerations. Unlike cancer, where early detection often leads to standardized interventions with favorable risk profiles, the management of bAVM is complex; as highlighted by the ARUBA trial, the short-term morbidity of surgical or endovascular intervention can exceed the short-term risk of bAVM rupture.^43^ Furthermore, identifying an “untreatable” lesion through population screening could lead to life-altering psychological distress without a clear path to risk reduction. However, this must be weighed against the devastating natural history of bAVM; while treatment carries risks, modern multimodal management including stereotactic radiosurgery can achieve safe obliteration rates of up to 80-90% for small-to-medium-sized lesions.^44^ Ultimately, the possibility of psychological harm or treatment-related injury must be balanced against the potential benefit of identifying and neutralizing a clinically silent pathology before a catastrophic hemorrhage occurs.

A unique aspect of our study worthy of some discussion is the combination of common and rare genetic variant risk for bAVM prediction. A byproduct of this approach is that we are able to report the prevalence of bAVM among individuals with rare pathogenic variants in bAVM-associated genes (*ENG*, *ACVRL1*, *SMAD4*, *GDF2, RASA1,* and *EPHB4*) ascertained through an entirely genotype-first methodology. Prior studies reporting bAVM rates in patients with pathogenic bAVM-associated variants have relied on phenotype-first or familial-variant-first ascertainment, where patients (and bAVM rate) are identified based on the presence of symptoms or based on a family history of the disease.^19–21^ Our study, including patients on the basis of genotype alone, found an overall bAVM rate of 4.7%. The bAVMs identified in this cohort were entirely confined to patients with pathogenic variants in two genes – *ENG* and *ACVRL1*, both causal of hereditary hemorrhagic telangiectasia. bAVMs were identified in 19% (3/16) of individuals with *ENG* pathogenic variants and in 3% (1/32) of patients with *ACVRL1* pathogenic variants – both on the upper end of what has been reported in the literature for patients ascertained through phenotype first approaches (10-17% for *ENG* and 1-4% for *ACVRL1*).^19^ We suspect that the high rate of bAVM in *ENG* and *ACVRL1* patients may be partly explained by ascertainment bias inherent to PMBB as a biobank within a tertiary care health system, where participants often present with higher disease burden. This hypothesis is supported by the fact that this overall PMBB study population had a 10-fold higher prevalence of bAVM (0.1%) than the commonly accepted estimate of 0.01%.^17^ However, at the same time, we observed a 0% prevalence of bAVM among carriers of *RASA1* (0/5) and *EPHB4* (0/20) pathogenic variants. This contrasts with the reported rates of 10% for RASA1 and 3% for *EPHB4*,^21^ suggesting that penetrance for bAVM conferred by these variants may be lower when identified in a genotype-first fashion. However, we note that the overall small sample size of patients with variants in either of these genes precludes us from making precise estimates of bAVM risk.

The performance of BAVM-PRS varied across ancestry groups, which was expected given that the score was derived from a European-ancestry GWAS (**Figure 2**). This likely reflects known cross-population differences in linkage disequilibrium structure and allele frequency distribution.^45–48^ While overall case separation (as measured by AUC) in the African-ancestry cohort was lower, the BAVM-PRS’s ability to identify high-risk individuals at extreme tails was preserved. Our findings also suggest that clinical deployment would benefit from ancestry-specific thresholds: an optimized cutoff of BAVM-PRS >2.8 in the African-ancestry cohort identified a high-risk subset with 3.0% prevalence (17.8-fold enrichment), whereas a threshold of BAVM-PRS >2.0 in the European-ancestry cohort yielded a 0.22% prevalence (3.1-fold enrichment). The ultimate utility of polygenic risk scoring in bAVM will require additional larger, multi-ethnic discovery GWAS to capture ancestry-specific risk.

Another interesting finding in our work was that biologically constraining our PRS framework to functionally relevant gene pathways did not degrade model performance, but rather appeared to improve it. Each of the variants included in our final BAVM-PRS had been mapped to genes whose role in bAVM development and vascular signaling had been demonstrated in functional studies prior to their identification in the source GWAS.^24^ Comparing performance of our biologically restricted model to a PRS construction including all GWAS-significant SNPs, we found that the all-SNP BAVM-PRS-All model had a lower AUC (0.682) and, surprisingly, weaker directional association with bAVM within both the European-and African-ancestry subpopulations. We hypothesize that the All-SNP model may be incorporating population-specific noise within SNPs that did not translate across cohorts, while the BAVM-PRS’s focus on biologically relevant pathways may have allowed greater portability of its risk signal. This approach is supported by recent evidence demonstrating that prioritizing variants in functional or tissue-specific regulatory elements significantly enhances the trans-population portability of polygenic risk scores.^31,32^

Our study has several limitations. While misclassification of bAVM cases was minimized using the strict criteria of chart review with radiographic confirmation of bAVM and exclusion of individuals with ICD-10 code Q28.2 but without imaging for review, there are almost certainly participants within the control group that we defined who have undiagnosed or non-imaged bAVM. This is evidenced by the fact that 2 of the bAVMs identified during chart review of the Mendelian cohort did not have an associated brain AVM ICD-10 code. A major limitation of polygenic risk scores is that they do not perform well on an individual basis.^25^ This drove our approach towards population risk stratification for directed screening rather than individual-level interventions. Additionally, as expected, our cohort of patients with rare Mendelian bAVM variants was relatively small (n=86), limiting our ability to accurately determine the true prevalence of bAVM in this group. Lastly, as a tertiary care biobank, the PMBB population carries an inherently higher degree of clinical acuity and diagnostic workups. The high baseline bAVM prevalence (0.1% in PMBB vs 0.01% in the general population) suggests that our calculated NNS values may be lower than what would be observed in a standard primary care or community-based setting. A community-based validation of the two-step genetic screening model would be a mandatory next step.

Overall, this study demonstrates that a two-step genetic risk model-combining Mendelian variant screening with a novel, biologically-constrained bAVM polygenic risk score (BAVM-PRS)-can identify individuals at high risk for brain arteriovenous malformations, with a number needed to screen of 122, comparable to established screening protocols for breast and lung cancer. While germline genetic risk represents only one component of bAVM’s multifaceted pathophysiology, our model provides a starting point for targeted MRI screening in a disease where the initial presentation in undiagnosed individuals is frequently a catastrophic hemorrhage. The broader implementation of a refined version of this approach can fundamentally shift our clinical approach to neurovascular disease: from reactionary interventions to a proactive, life-saving campaign of early detection.

## Data Availability

Individual-level data, aside from what has been included in the manuscript and supplemental materials, cannot be shared due to patient privacy concerns and as patient participants have not been broadly consented for dissemination of individual-level data, except where otherwise noted. The PennMedicine BioBank (PMBB) data used in this study were generated previously, and PMBB data are available to researchers through collaboration with PMBB via the PMBB website at https://pmbb.med.upenn.edu/investigators.php. Non-identifiable summary-level data in the present study are available upon reasonable request to the authors.

## Acknowledgements

We acknowledge the Penn Medicine BioBank (PMBB) for providing data and thank the patient-participants of Penn Medicine who consented to participate in this research program. We would also like to thank the Penn Medicine BioBank team and Regeneron Genetics Center for providing genetic variant data for analysis. The PMBB is approved under IRB protocol# 813913 and is supported by the Perelman School of Medicine at the University of Pennsylvania, by a gift from the Smilow family, and by the National Center for Advancing Translational Sciences of the National Institutes of Health under CTSA award number UL1TR001878. This specific study was approved under IRB protocol #858694.

## Author Contributions

P.D.P. and T.G.D. carried out the conceptualization and design of the study. P.D.P. performed the formal analysis and data preparation. P.D.P. and T.G.D. verified the analytical methods. M.R.G., A.K., S.M.D., S.T., J.K.B., P.J., and T.G.D. supervised the project. P.D.P. took the lead in writing the manuscript. All authors (P.D.P., S.S., M.R.G., A.K., S.M.D., S.T., J.K.B., P.J., and T.G.D.) provided critical feedback and helped shape the final manuscript.

## Data and Code Availability

Individual-level data, aside from what has been included in the manuscript and supplemental materials, cannot be shared due to patient privacy concerns and as patient participants have not been broadly consented for dissemination of individual-level data, except where otherwise noted. The PennMedicine BioBank (PMBB) data used in this study were generated previously, and PMBB data are available to researchers through collaboration with PMBB via the PMBB website at https://pmbb.med.upenn.edu/investigators.php.

## Supplemental Information

**Table S1:**
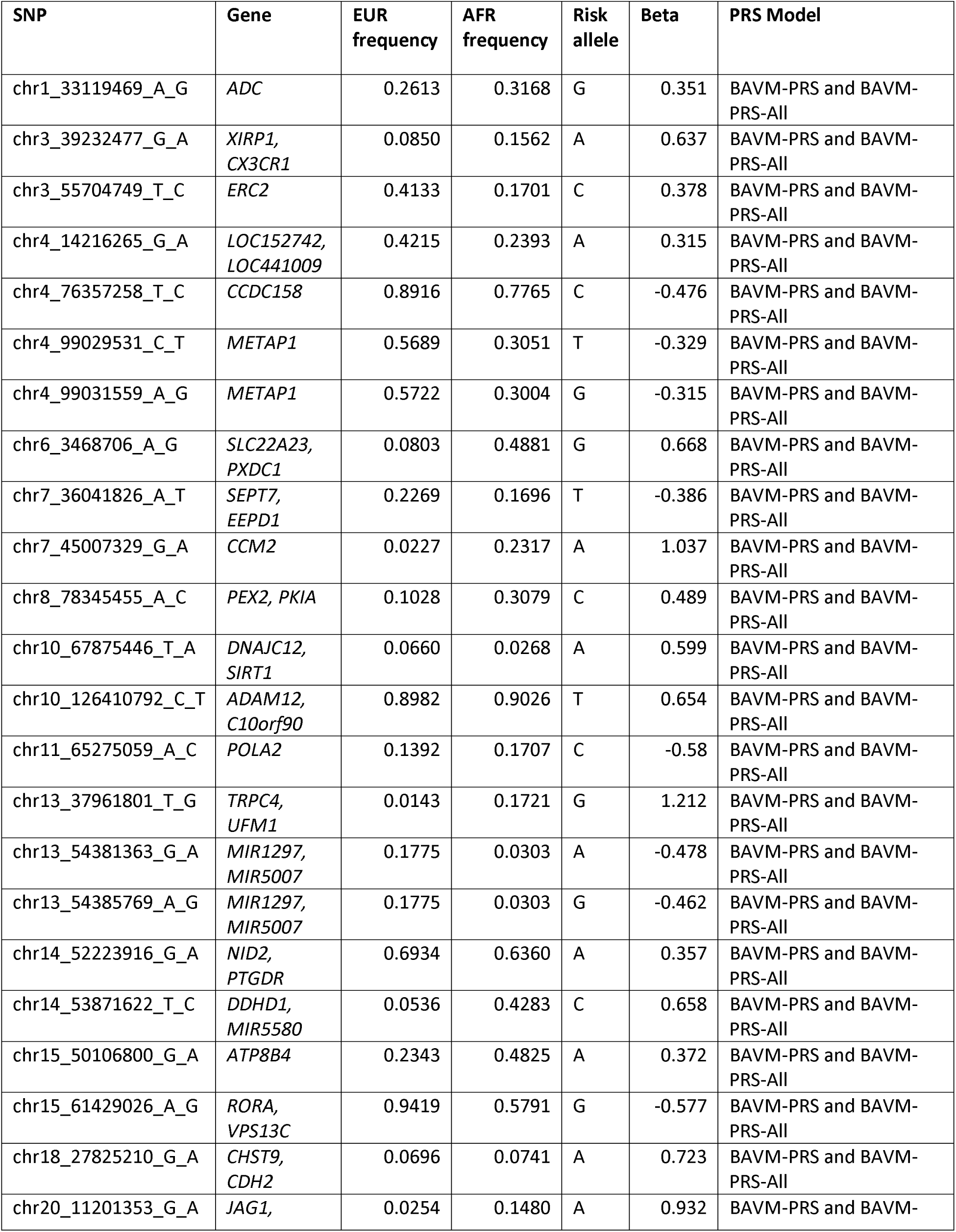
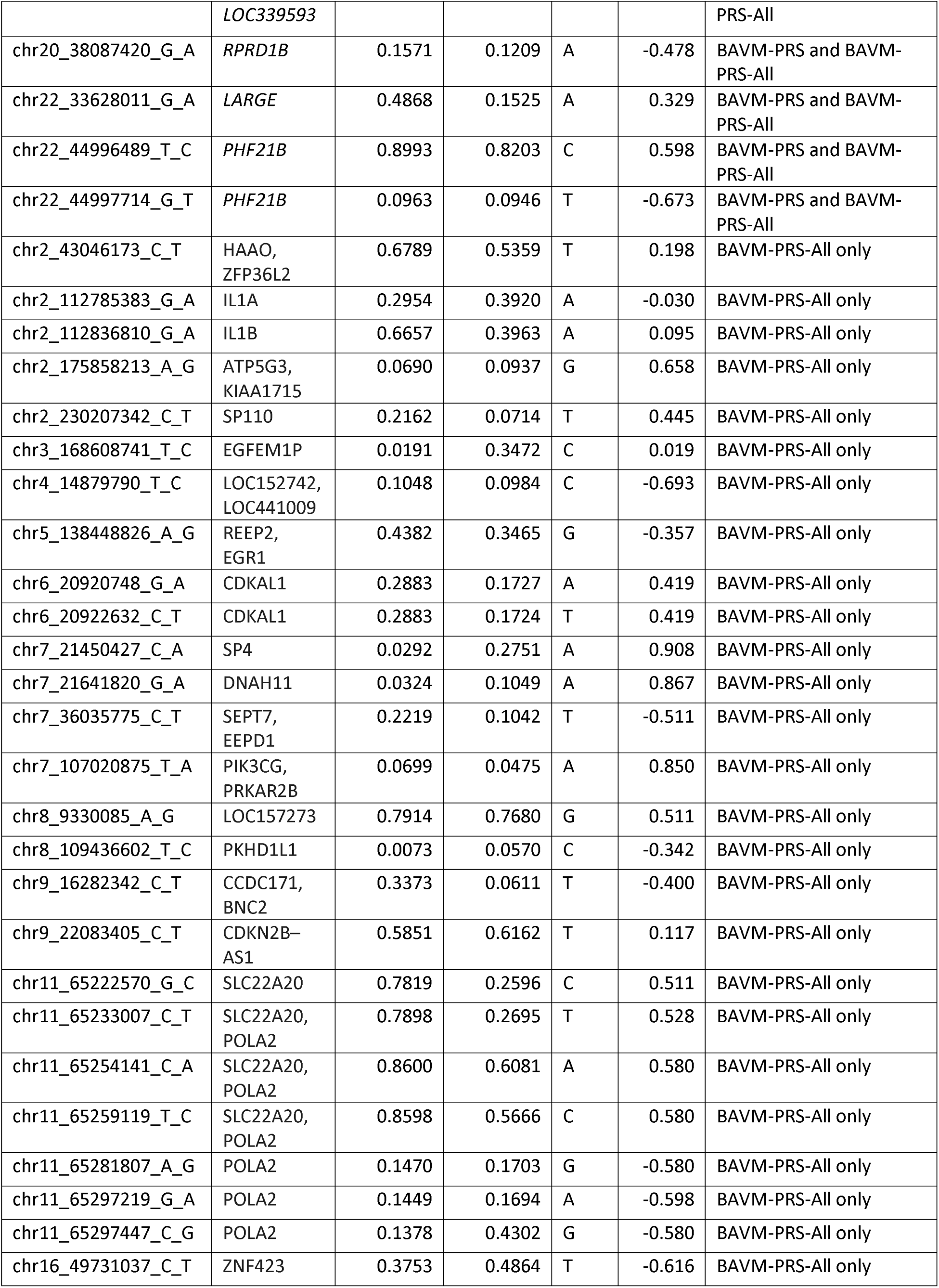
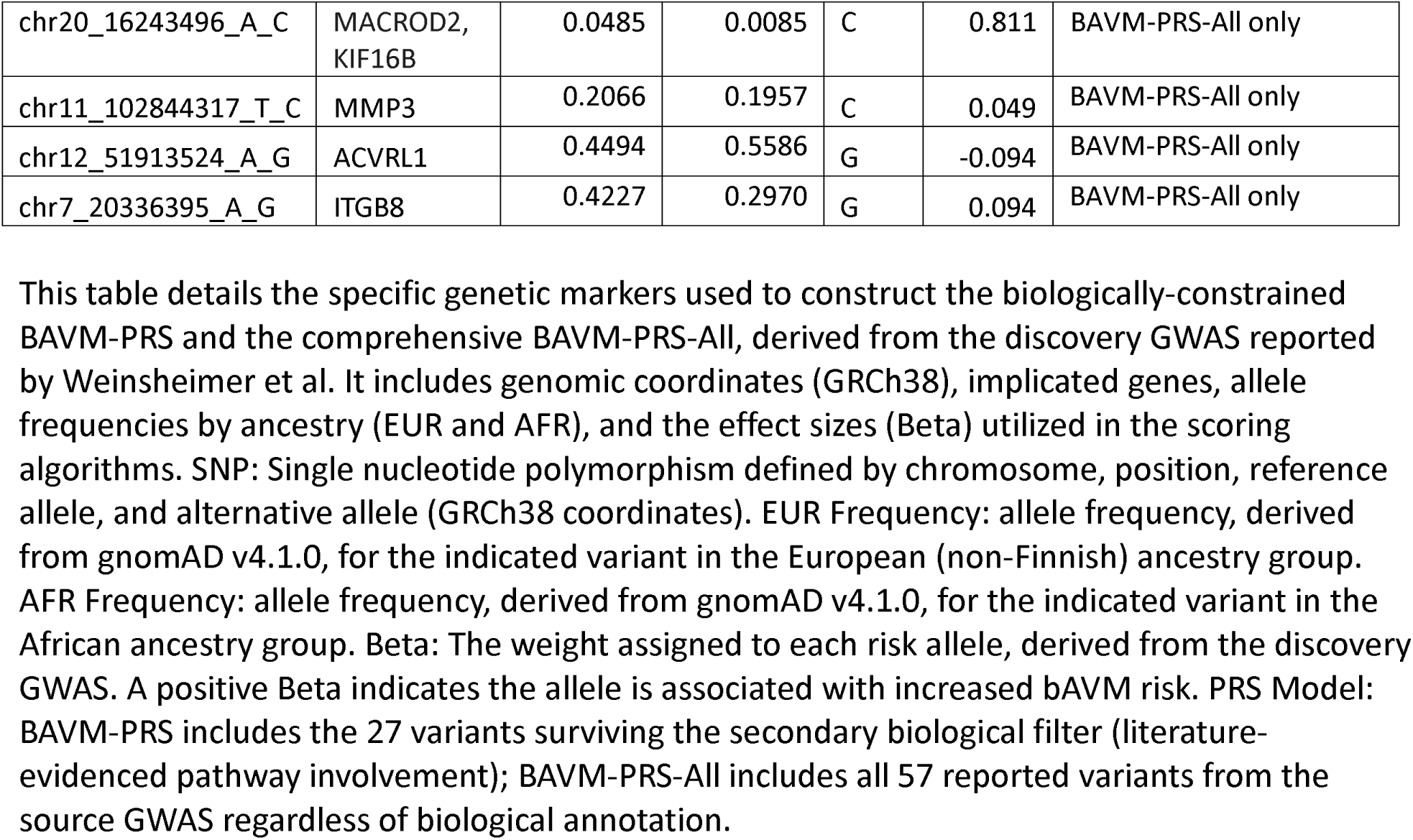
PRS Score Components.

---

**Figure S1:**
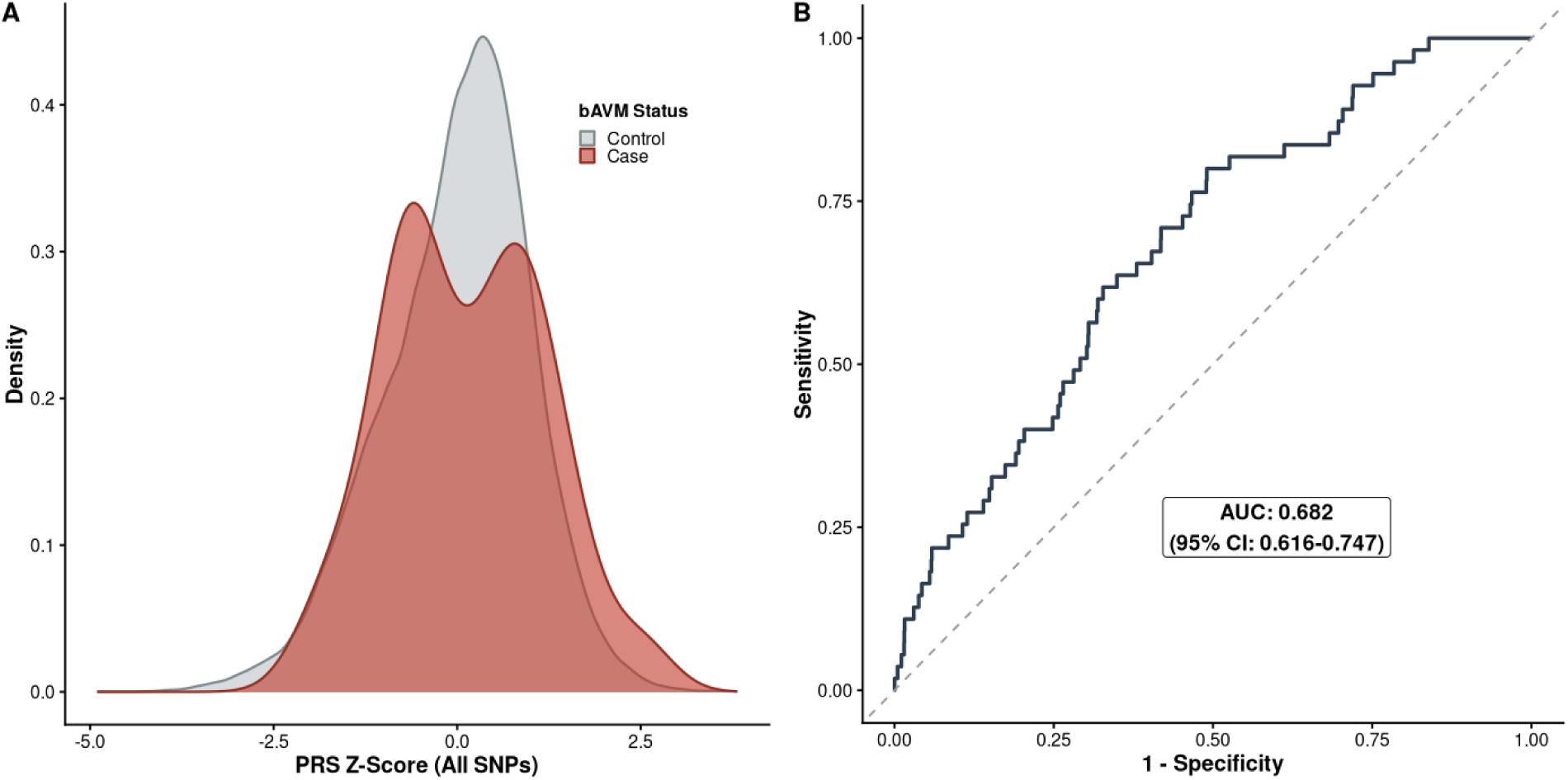
Performance of the All-Inclusive BAVM-PRS-All Without Biological Restriction in the Non-Mendelian Cohort. This figure evaluates the diagnostic utility of BAVM-PRS-All, a PRS constructed using all available SNPs from the discovery GWAS (Weinsheimer et al.), without limiting the model to biologically relevant or functionally informed variants. (A) Density Distribution of BAVM-PRS-All scores (represented as Z-score) is shown for confirmed bAVM cases (red, n = 55) and controls (grey, n = 56,191) within the non-Mendelian cohort. There is no significant shift in the median PRS for cases compared to controls (median [IQR] 0.072 [-0.643, 0.905] vs. 0.107 [-0.619, 0.681], p=0.613). Unlike the primary BAVM-PRS, the all-inclusive model demonstrates a notable bimodal distribution among bAVM cases. This lack of a clear, single directional shift suggests that the inclusion of non-biologically informed SNPs introduces noise that detracts from a clean directional association with disease status. (B) Receiver operating characteristic (ROC) curve illustrating the discriminatory performance of BAVM-PRS-All for identifying brain arteriovenous malformations in the non-Mendelian PMBB population. The BAVM-PRS-All achieved an area under the curve (AUC) of 0.682 (95% CI: 0.616-0.747). While the AUC is numerically similar to the primary BAVM-PRS (0.694), the all-inclusive, BAVM-PRS-All model lacked a significant directional association with bAVM status in logistic regression analysis (**Table 3**), likely due to the inconsistent distribution observed in the case cohort. These results support the use of a biologically informed PRS over an all-inclusive model. Restricting the PRS to variants within genes relevant to vascular stability and development (e.g., TGF-beta signaling, endothelial function) results in a more robust and directionally consistent association with bAVM diagnosis. Abbreviations: AUC, area under the curve; bAVM, brain arteriovenous malformation; CI, confidence interval; PMBB, Penn Medicine BioBank; PRS, polygenic risk score; IQR, interquartile range.

**Figure S2:**
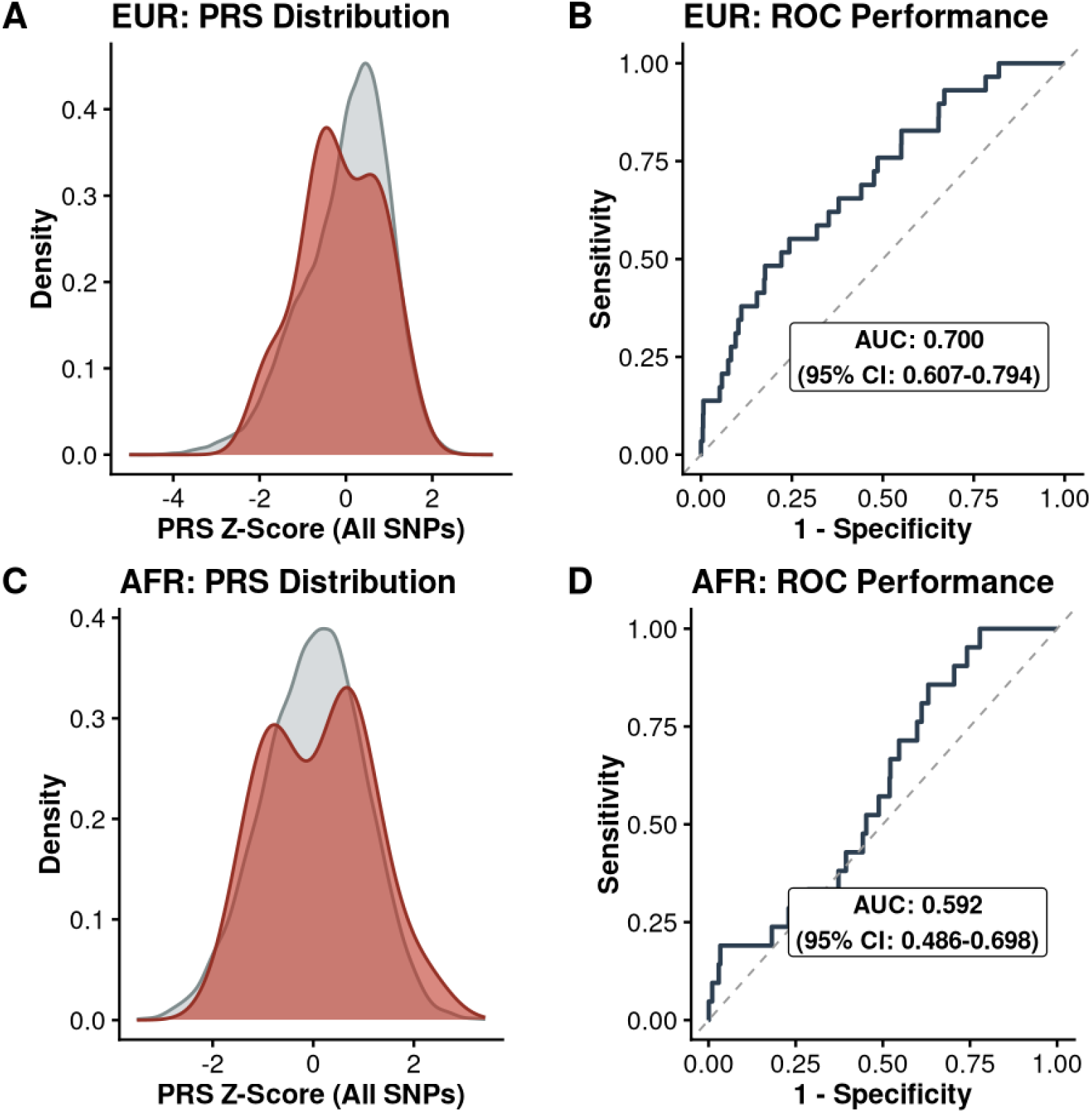
Ancestry-Stratified Performance of BAVM-PRS-All in the Non-Mendelian Cohort. This figure displays the performance of the non-restricted BAVM-PRS-All across European and African ancestral subgroups. (A) Density Distribution of BAVM-PRS-All scores (represented as Z-score) is shown for confirmed bAVM cases (red, n = 29) and controls (grey, n = 41,546) within the non-Mendelian cohort specifically for individuals of European GIA. (B) Receiver operating characteristic (ROC) curve illustrating the discriminatory performance of BAVM-PRS-All for identifying brain arteriovenous malformations in the non-Mendelian PMBB population for individuals of European GIA. The BAVM-PRS-All achieved an area under the curve (AUC) of 0.700 (95% CI: 0.607–0.794), representing modest discriminating ability. (C) Density Distribution of BAVM-PRS-All scores (represented as Z-score) is shown for confirmed bAVM cases (red, n = 21) and controls (grey, n = 11,988) within the non-Mendelian cohort specifically for individuals of African GIA. (D) Receiver operating characteristic (ROC) curve illustrating the discriminatory performance of BAVM-PRS-All for identifying brain arteriovenous malformations in the non-Mendelian PMBB population for individuals of African GIA. The BAVM-PRS-All achieved an AUC of 0.592 (95% CI: 0.486–0.698). While the AUC values remain numerically comparable to the ancestry-specific performance of the biologically informed PRS (Figure 2), the all-inclusive model’s bimodal case distribution and lack of significant directional association (**Table 3**) limit its clinical utility for risk stratification. Abbreviations: AFR, African; AUC, area under the curve; bAVM, brain arteriovenous malformation; CI, confidence interval; EUR, European; PRS, polygenic risk score.

## Supplemental Note 1: List of PMBB contributors

### PMBB Leadership Team

Daniel J. Rader, M.D., Marylyn D. Ritchie, Ph.D.

#### Contribution

All authors contributed to securing funding, study design and oversight. All authors reviewed the final version of the manuscript.

### Patient Recruitment and Regulatory Oversight

JoEllen Weaver, Nawar Naseer, Ph.D., M.P.H., Giorgio Sirugo, M.D., Ph.D., Afiya Poindexter, Yi-An Ko, Ph.D., Kyle P. Nerz

#### Contributions

JW manages patient recruitment and regulatory oversight of study. NN manages participant engagement, assists with regulatory oversight, and researcher access. GS assists with researcher access. AP, YK, KPN perform recruitment and enrollment of study participants.

### Lab Operations

JoEllen Weaver, Meghan Livingstone, Fred Vadivieso, Stephanie DerOhannessian, Teo Tran, Julia Stephanowski, Salma Santos, Ned Haubein, Ph.D., Joseph Dunn

#### Contribution

JW, ML, FV, SD conduct oversight of lab operations. ML, FV, AK, SD, TT, JS, SS perform sample processing. NH, JD are responsible for sample tracking and the laboratory information management system.

### Clinical Informatics

Anurag Verma, Ph.D., Colleen Morse Kripke, M.S. DPT, MSA, Marjorie Risman, M.S., Renae Judy, B.S., Colin Wollack, M.S.

#### Contribution

All authors contributed to the development and validation of clinical phenotypes used to identify study participants and (when applicable) controls.

### Genome Informatics

Anurag Verma, Ph.D., Shefali S. Verma, Ph.D., Scott Damrauer, M.D., Yuki Bradford, M.S., Scott Dudek, M.S., Theodore Drivas, M.D., Ph.D.,

#### Contribution

AV, SSV, and SD are responsible for the analysis, design, and infrastructure needed to quality control genotype and exome data. YB performs the analysis. TD and AV provide variant and gene annotations and their functional interpretation of variants.

## Notes

### Competing Interest Statement

Dr. Jabbour is a consultant for Medtronic, MicroVention, Balt and Cerus Endovascular. Dr. Tjoumakaris is a consultant for MicroVention. Dr. Gooch is a consultant for Stryker.

### Author Declarations

IRB of University of Pennsylvania gave ethical approval for this work.

